# Integrating cognitive, linguistic and acoustic features to identify individuals with cognitive impairment: a proof-of-concept study

**DOI:** 10.64898/2026.08.25.26361344

**Authors:** Melody M.Y. Chan, Gail A. Robinson

## Abstract

Early identification of cognitive impairment remains challenging in settings where comprehensive cognitive and clinical assessments are not available. Acoustic and linguistic features in naturalistic speech may serve as useful behavioural markers of cognitive impairment, but the value of integrating these measures with cognitive assessment remains unclear. We tested whether combining acoustic and linguistic features from one-minute speech samples with multi-domain cognitive assessment (spanning attention, language, memory and executive functions) improves classification of cognitively unimpaired individuals from those with amnestic mild cognitive impairment or early-stage Alzheimer’s Disease. Across multiple machine learning models, combining cognitive, acoustic and linguistic features yielded significantly better classification performance than models using cognitive or speech features alone (area under the curve = 0.96-0.98, both comparisons p < .05). This proof-of-concept study reveals that integrating speech-based measures with cognitive testing may improve identification of cognitive impairment, supporting the development of accessible and scalable multimodal screening tools for primary care.

## Introduction

Dementia is a clinical syndrome characterised by progressive difficulties in memory, language, and behaviour that gradually interferes with activities of daily living [1]. Despite continued research efforts, it remains a global health challenge with considerable socioeconomic consequences. Alzheimer’s disease (AD) and other dementias are projected to cost the global economy approximately INT$14.5 trillion between 2020 and 2050 [2]. Although there is currently no cure for dementia, earlier identification may enable timely intervention, care planning, and access to support while functional independence is relatively preserved [3]. Specifically, earlier diagnosis is associated with slower disease progression [4], reduced risk of mortality [5], and lower caregiver emotional burden [6]. Thus, optimising ways to support early identification remains an important area of research.

Primary care is often the first point of contact for individuals with concerns about cognitive decline [1]. It plays an important role in determining whether further specialist assessments, e.g., blood-based biomarker tests [7], are warranted. In many primary care settings, this initial assessment relies heavily on brief cognitive screening tools, which assess domains including memory, language, executive function, attention, and visuospatial abilities. Cognitive screening tools are increasingly accessible, with digital versions and app-based assessments now available in many countries [8, 9]. However, an important question is whether cognitive testing alone is sufficient. Cognitive measures capture performance under specific testing conditions and may not encompass the broader behavioural manifestations of early cognitive impairment. Indeed, a systematic review found that although many brief cognitive tests can distinguish clinical Alzheimer-type dementia from healthy cognition with reasonable accuracy, their ability to discriminate earlier stages of impairment from normal cognition is more limited [10].

Speech has recently emerged as a potentially useful source of information for detecting cognitive decline [11]. Speech contains a wide range of linguistic and acoustic features that may capture broader behavioural changes associated with cognitive impairment. For example, lexical-semantic characteristics of speech have been associated with beta-amyloid status [11], while speech features during memory recall relate to tau burden in early adulthood [12], suggesting that speech may reflect subtle AD-related behavioural changes. Importantly, these features can be extracted from brief, structured speech samples, providing a potentially scalable and low-burden source of behavioural information. However, demonstrating that speech features can distinguish individuals with cognitive impairment from cognitively healthy adults does not establish whether they capture information beyond that reflected in cognitive screening. If speech captures subtle behavioural manifestations of cognitive impairment that are not fully reflected in structured cognitive testing, integrating the two sources of information may improve classification relative to either alone.

The present proof-of-concept study examined whether integrating speech-derived linguistic and acoustic measures with multi-domain cognitive assessment improves classification of individuals with amnestic mild cognitive impairment (aMCI) and early AD relative to either modality (i.e., cognitive/speech) alone. Using a sample of 114 participants, we applied multiple machine learning algorithms to compare the classification performance of cognitive, linguistic, and acoustic measures, both separately and in combination, for distinguishing cognitive healthy individuals from those with aMCI/AD. We also examined the relative performance of the two unimodal approaches to characterise the contribution of each source of information to classification. More broadly, this study evaluates the potential of integrating established cognitive assessment with scalable speech-based measures to support more effective and accessible early identification of cognitive impairment.

## Methods

### Participant and procedure

Participants were drawn from the onsite cohort of the Prospective Imaging Study of Ageing: Genes, Brain and Behaviour (PISA). This study, targeting individuals aged 40-80, was designed to characterise the biological and behavioural markers associated with the onset of AD. The PISA study protocol was approved by the Human Research Ethics Committees of QIMR Berghofer Medical Research Institute and The University of Queensland, which was reported in Lupton, Robinson [13]. In brief, PISA comprised a cognitively healthy cohort recruited from an existing QIMR Berghofer database, and a clinical cohort of individuals with aMCI or mild AD (Mini-Mental State Examination >20 and Clinical Dementia Rating of 0.5 or 1.0). Participants underwent extensive phenotyping, including neuropsychological assessment, multimodal neuroimaging [magnetic resonance imaging (MRI) and amyloid positron emission topography (PET)], blood sampling, and speech sample collection. The clinical status for each participant was confirmed by a multidisciplinary team who were blinded to participant’s amyloid PET results. For this study, 114 participants (mean age: 61.9 years, s.d.: 7.68 years) with available neuropsychological data and speech audio recordings and transcripts were included (**Figure 1a**). Fifty-six participants were classified as cognitively healthy, and the remaining 58 participants were classified as cognitively impaired (aMCI n = 25; mild AD n = 33) through clinical consensus. Their cognitive test scores were used directly, while speech audio recordings and transcripts underwent additional acoustic and linguistic feature extraction, detailed below.

**Figure 1:**
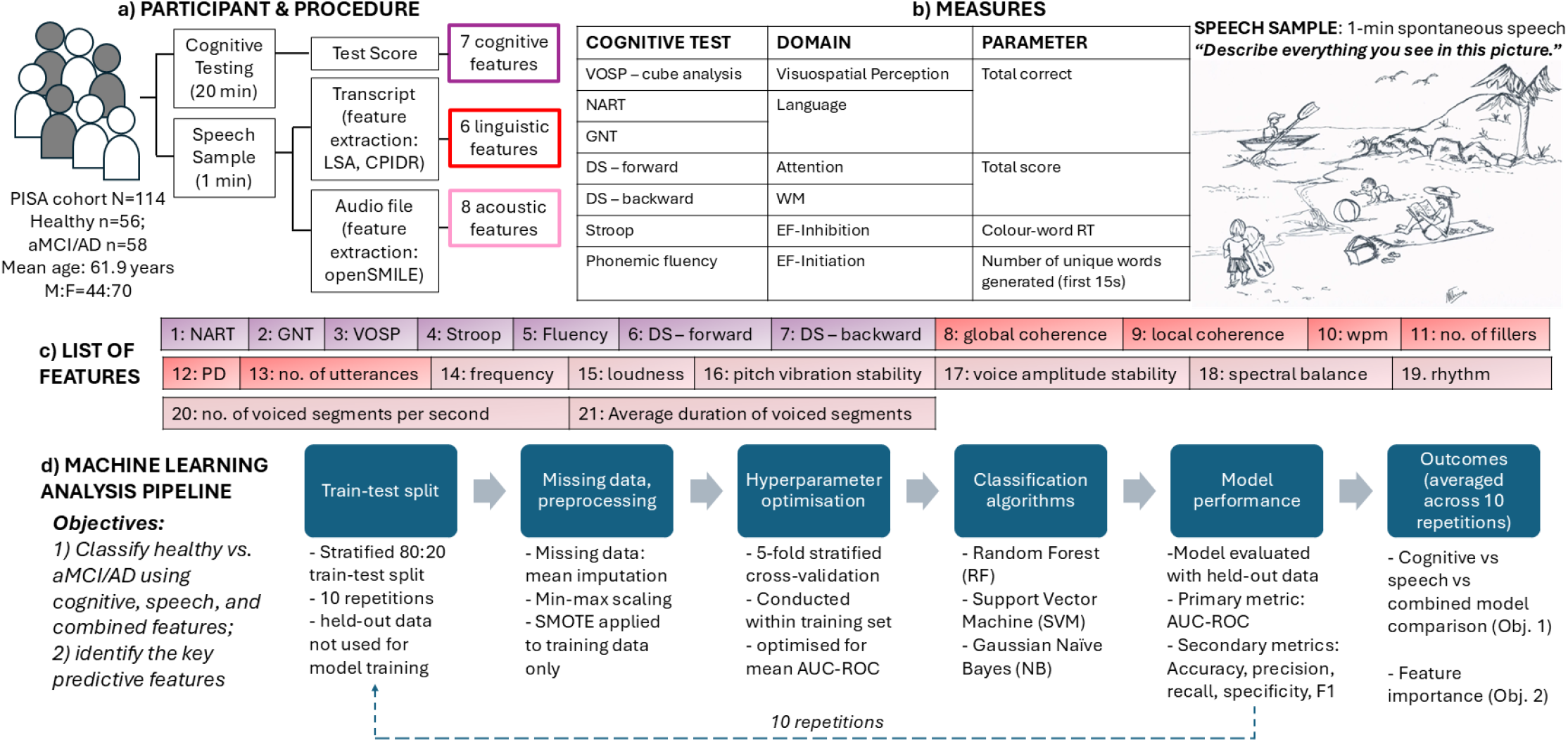
Study design and machine learning analysis pipeline. Cognitive and speech data from 114 participants were analysed. All of them underwent cognitive testing, with a speech sample also collected during the same testing session. Neuropsychologists administered the test and collected speech samples in a quiet testing room. After data collection, tests were scored by a neuropsychologist, and audio-recorded speech samples were transcribed by trained research personnel. While test scores were directly used as cognitive features, transcripts and speech audio recordings were further processed to generate linguistic and acoustic features (**panel a**). Cognitive and speech measures used for probing cognitive and language functions are shown in **panel b**. The full set of cognitive, linguistic and acoustic features is shown in **panel c**. The number corresponds to feature numbers shown in **Figure 2c** (feature importance analysis results). **Panel d** outlines the machine learning pipeline used in this study. The data were first split into training and test sets, and preprocessed for optimising machine learning model performance. Multiple algorithms were trained to classify healthy and cognitively impaired participants based on the extracted features, and the classification performance of each of the trained models were evaluated using the independent test sets. For each model, feature importance analyses were conducted to identify key features supporting classification. *Abbreviations:* LSA=latent semantic analysis; CPIDR=Computerized Propositional Idea Density Rater; VOSP=Visual Object and Space Perception Battery; NART=National Adult Reading Test; GNT=graded naming test; DS=digit span; WM=working memory; EF=executive function; RT=response time; wpm=words per minute; PD=propositional density; Obj.= objective.

### Measures and feature extraction

The neuropsychological tests and spontaneous speech task used to derive the cognitive, linguistic, and acoustic features are shown in **Figure 1b**.

#### Cognitive features

Seven neuropsychological measures were selected to tap core cognitive domains relevant to cognitive impairment (**Figure 1b**), including language [National Adult Reading Test (NART; [14]): word reading; Graded Naming Test (GNT; [15]): naming], visuospatial processing [Cube Analysis - Visual Object and Space Perception Battery (VOSP; [16])], executive function (Stroop [17]: inhibition; Phonemic fluency [18]: response initiation), auditory attention (digit span (DS) – forward [19]), and working memory (DS – backward [19]). Tests were administered and scored by neuropsychologists (AC, CS, EG, PT, KH, MB, and TR) supervised by a consultant clinical neuropsychologist (GR). Raw scores were used in the analyses described below.

#### Speech features

Participants completed a complex scene description task [20]. They were asked to describe everything they could see in a line drawing in one minute (**Figure 1b**). They were given one minute to describe everything they saw in a line drawing. Responses were audio recorded and transcribed by a trained research personnel, with transcripts independently checked by a neuropsychologist. Transcripts were used to derive linguistic features, while the corresponding audio recordings were used to extract acoustic features.

#### Global coherence (GC) and local coherence (LC)

GC and LC were calculated using the automated coherence analysis procedure developed by Hoffman, Loginova [21]. This approach uses latent semantic analysis (LSA) to represent the semantic content of speech within a high-dimensional vector space. GC is quantified as the similarity between each speech window and a composite semantic representation of responses to the same prompt, reflecting the extent to which the participant remains semantically focused on the target topic. LC is quantified as the cosine similarity between the semantic vector for each window and that of the immediately preceding window, reflecting the extent to which successive portions of speech remain semantically related. Each transcript was divided into overlapping 20-word windows. The analysis proceeded across the transcript using a moving window shifted by one word at a time. GC and LC values were averaged across all available windows for each participant’s response and multiplied by 100 for ease of interpretation. Higher values indicate greater GC/LC.

#### Other linguistic features

Number of fillers, words per minute (wpm), propositional density (PD), and number of utterances were derived from manually transcribed responses. During transcription, trained neuropsychologists identified and counted fillers (e.g., uh, um, mm, ah, and related variants), segmented responses to utterances (i.e., a continuous segment of speech bounded by a perceptible pause or the end of a spoken unit), and marked proposition boundaries using slash delimiters according to the Computerized Propositional Idea Density Rater (CPIDR) criteria described in Brown, Snodgrass [22]. Fillers were excluded from the wpm and PD calculations. PD was calculated as the number of propositions divided by the total number of words in the response.

#### Acoustic features

Eight acoustic measures were included, comprising fundamental frequency, loudness, pitch vibration stability (i.e., jitter), voice amplitude stability (i.e., shimmer), spectral balance (i.e., Hammarberg index), speech rhythm (i.e., loudness peak rate), number of voiced segments per second (i.e., voiced segment rate), and average duration of voiced segments (i.e., mean voiced segment length). These measures were reported to be relevant to AD [11] and were therefore selected to characterise different aspects in speech production. Frequency and loudness captured fundamental spectral and intensity-related properties of the speech signal. Jitter and shimmer characterised cycle-to-cycle variability in frequency and amplitude, respectively. The Hammarberg index represented spectral energy distribution. Loudness peak rate, voiced segment rate, and mean voiced segment length characterised temporal and prosodic aspects of continuous speech production. Acoustic features were extracted from the speech recordings using openSMILE [23] with the Geneva Minimalistic Acoustic Parameter Set (GeMAPS; [24]).

### Machine learning pipeline

The feature extraction procedure described above yielded 21 prespecified cognitive, linguistic and acoustic features, summarised in **Figure 1c**. These features were grouped into three feature sets: cognitive features (7 features), speech features comprising acoustic and linguistic measures (14 features), and a combined feature set (21 features). Classification performance of these feature sets was compared across multiple machine learning models. Analyses were performed using scikit-learn via Python 3.12. Three complementary supervised machine learning algorithms were evaluated: support vector machine (SVM), random forest (RF), and Gaussian naïve Bayes (NB). These algorithms were selected because they represent distinct approaches to classification and make different assumptions regarding the relationship between predictors and cognitive status. Applying the same feature-set comparisons across multiple algorithms also allowed assessment of whether the incremental value of combining cognitive and speech-derived information was robust to the choice of classifier. A summary of the pipeline is shown in **Figure 1d**. For further details, see *Supplementary methods*.

#### Model comparison

The primary analysis examined whether integrating cognitive- and speech-derived features improved classification performance relative to either feature set alone. Within each classifier, the area under the receiver operating characteristic curve (AUC) was compared across the same ten repeated train-test partitions for two paired contrasts: combined vs. speech, combined vs. cognitive. For each repetition, the difference in AUC (ΔAUC) was calculated and averaged across repetitions, with positive values indicating better performance of the combined feature set. Paired samples t-tests were conducted across the repeated partitions to compare AUC between feature sets. As a secondary analysis, speech-only and cognitive-only models were also compared to characterise the relative discriminatory performance of the two unimodal feature sets.

#### Feature importance

Feature importance analyses were conducted on the feature set yielded the highest AUC to identify the predictors contributing most strongly to classification. Model-specific importance measures were used: impurity-based importance for random forest, absolute coefficients for SVM, and absolute class-specific mean differences for NB. Importance values were normalised within each model and averaged across the ten repetitions. This provided a relative importance measure within each model. Features consistently ranked among the most important predictors across classifiers were identified as robust contributors to classification, as their importance was less dependent on the characteristics of any single modelling approach.

## Results

Across all three classifiers, the combined cognitive-speech feature set achieved the highest mean AUC, followed by speech-derived and cognitive-only features (**Figure 2a**). Mean AUCs ranged from .859-.897 for cognitive features, .933-.956 for speech-derived features, and .959-.986 for combined features. The highest AUC was observed for the combined Naïve Bayes model (.986). Other performance measures, including accuracy, sensitivity, specificity and F1 score, are reported in *Supplementary results*.

**Figure 2:**
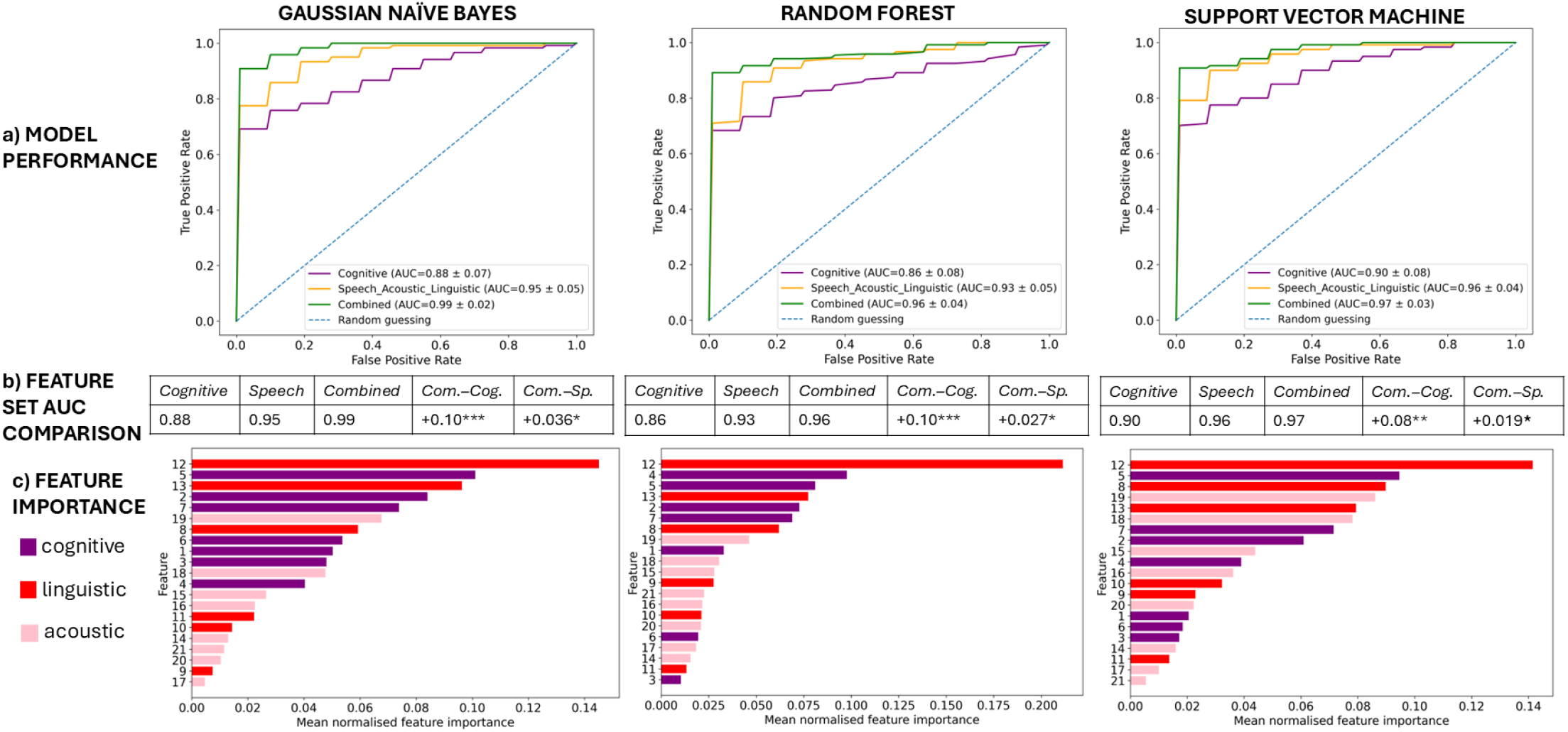
Main results – classification model performance and feature importance. **Panel a** shows the receiver operating characteristic (ROC) curves for the gaussian naïve bayes, random forest, and support vector machine models across 10 repeated train-test splits. For each classification model, curves compare the discriminative performance with cognitive, speech (i.e., acoustic and linguistic features) and combined (i.e., cognitive, acoustic and linguistic features) feature sets. Discriminative performance was quantified using the area under the ROC curve (AUC), reported as mean +/-standard deviation across repeated splits. The dashed diagonal line represents performance expected from random classification. **Panel b** presents the AUC values across the 10 repeated train-test splits for each feature set and classification model. Prespecified contrasts compared the combined model with the speech-only and cognitive-only models (i.e., Com.-Sp. And Com.-Cog., respectively). Statistical significance was assessed using planned paired-samples t-tests on corresponding split-level AUC values across the 10 repeated splits. **Panel c** shows the feature importance for each classification algorithm using the combined feature set. Feature importance was quantified using model-specific measures, normalised within each model, and averaged across the 10 repeated splits. Some features showed consistently high relative importance across models. Feature numbers correspond to the numbered feature key shown in **Figure 1c**. *Significance levels:* *p<.05, **p<.01. ***p<.001.

Paired-samples t-tests across matched train-test partitions (**Figure 2b**) showed that the combined feature set significantly outperformed cognitive features alone across all classifiers (NB: ΔAUC = .105, p < .001; RF: ΔAUC = .100, p = .002; SVM: ΔAUC = .078, p = .009). Combined models also significantly outperformed speech-derived features alone (NB: ΔAUC = .036, p = .019; RF: ΔAUC = .027, p = .014; SVM: ΔAUC = .019, p = .031). Thus, integrating cognitive and speech-derived features improved classification performance relative to either feature set alone. The improvement was larger relative to cognitive-only models than relative to speech-only models. The comparison between speech- and cognitive-only models are reported in *Supplementary results*.

Feature importance analyses on the combined cognitive-speech feature set showed that, highly ranked predictors in the combined models were distributed across cognitive, linguistic and acoustic domains (**Figure 2c**). Several individual features were consistently highly ranked across classifiers, including PD (feature 12), fluency (feature 5), number of utterances (feature 13), global coherence (feature 8), DS – backward (feature 7) and speech rhythm (feature 19), suggesting that their contribution to classification was not specific to a single modelling approach.

## Discussion

In this study, we investigated whether cognitive and speech-derived measures could be used to distinguish cognitively healthy individuals from those with aMCI/mild AD, and whether integrating these sources of information improved classification performance. Across three machine learning classifiers, speech-derived features provided stronger discrimination than cognitive measures alone, while models combining cognitive and speech features consistently achieved the highest AUC. Feature importance analyses further identified a group of cognitive and speech-derived measures that contributed consistently across classifiers. Together, these findings suggest that cognitive performance and characteristics of spontaneous speech provide partially distinct information about cognitive status (cognitively intact vs. impaired), and that their integration may offer a more informative behavioural profile of cognitive impairment than either source alone.

The key finding was that combining cognitive and speech-derived measures consistently improved discrimination across classifiers, as reflected by higher AUCs for the combined models. Cognitive testing captures performance under structured conditions, whereas spontaneous speech reflects how multiple cognitive processes are expressed during everyday language production. Importantly, both represent behavioural manifestations of cognitive functioning and, despite increasing interest in biological biomarkers of neurodegeneration, the present findings highlight that relatively simple behavioural measures continue to contain substantial information for detecting cognitive impairment. Rather than relying on a single cognitive score or speech marker, integrating information across these behavioural domains may provide a more complete representation of an individual’s cognitive profile.

Feature importance analyses provided further insight into why combining these measures improved classification. Several predictors were consistently highly ranked across classifiers, including propositional density, response initiation, number of utterances, global coherence and working memory, while speech-rhythm measures (i.e., loudness peak rate) were also highly influential in more than one classifier. Several of these measures have previously been examined individually in relation to age-related cognitive decline (e.g., global coherence [21]; propositional density [25]). The present findings extend this work by showing that their value may lie in their joint contribution to a multivariate behavioural profile. Rather than identifying a single optimal marker, the results suggest that classification benefits from integrating multimodal information. The consistency of several highly ranked features across different classifiers also indicates that their contribution was not restricted to the feature importance characteristics of a single modelling approach.

These findings have important implications for cognitive assessment in primary care and other low-resource settings. Cognitive screening and speech analysis have long been considered as separate approaches in detecting cognitive impairment. However, the present findings suggest that integrating information from both may be more informative than relying on either modality alone. Both sources of information are comparatively low burden and potentially amenable to digital administration. Unlike neuroimaging, cerebrospinal fluid analysis, or other specialist investigations, a brief cognitive assessment and a short speech sample could feasibly be collected using a digital platform with minimal equipment. Cognitive tasks could be scored automatically, while speech could be recorded, automatically transcribed, and processed to derive linguistic and acoustic measures without requiring extensive manual analysis. This creates the possibility of integrating cognitive and speech assessment within a single digital screening tool that could be used as an initial assessment before referral for more comprehensive evaluation. Such an approach may be particularly valuable in primary care, where time and access to specialist cognitive assessment can be limited, and could potentially extend assessment to remote or underserved populations. The present study does not establish the clinical utility of such a system, however; digital tool development and future validation studies in primary care settings would be required before implementation.

To our knowledge, this is the first study to demonstrate that the combined cognitive-speech profile outperforms its individual components in distinguishing cognitively healthy individuals from those with aMCI/mild AD. These findings have important implications for the future development of scalable early identification strategies for dementia, particularly integrated digital approaches that combine brief cognitive assessment with automated speech analysis. Yet, several limitations should be acknowledged. The relatively modest sample size and lack of external validation limit the generalisability of the findings. In addition, the cross-sectional design demonstrates classification of current cognitive status rather than prediction of future cognitive decline. Future studies should therefore validate these findings in larger, independent and longitudinal cohorts, including more diverse populations speaking different languages. Nonetheless, these findings provide proof-of-concept that combining brief cognitive measures with speech-derived features can improve identification of individuals with cognitive impairment. With further validation, it could be translated into scalable digital screening pathways for broader population-level applications.

## Data Availability

All data produced in the present study are available upon reasonable request to the authors.

## Acknowledgements

The authors wish to thank Amelia Ceslis, Chris Schumann, Kristina Horne, Emily Gibson, Priscilla Tjokrowijoto, and Toby Rheinhardt for contributing to participant testing, cognitive test scoring and speech data transcription. At the time of data collection, G.R. was supported by an Australian National Health and Medical Research Council (NHMRC) Boosting Dementia Research Leadership Fellowship (APP1135769).

