## Supplementary methods for "Integrating cognitive, linguistic and acoustic features to identify individuals with cognitive impairment: a proof-of-concept study"

#### *Machine learning algorithm parameters*

***Support vector machine (SVM).*** SVM classification was implemented using a linear kernel. The SVM identifies a decision boundary that maximises the margin separating observations from the two classes. A linear kernel was selected to provide a relatively parsimonious decision function given the sample size and number of predictors. The regularisation parameter, C, which controls the trade-off between maximising the classification margin and penalising misclassification within the training data, was optimised over values of 0.1, 1 and 10. Probability estimates were enabled to permit calculation of receiver operating characteristic curves and AUC.

***Random forest (RF).*** RF classification was used as an ensemble tree-based approach capable of accommodating nonlinear associations and interactions among predictors without requiring these relationships to be specified a priori. RF generates multiple decision trees using bootstrap samples of the training data and aggregates predictions across the resulting ensemble. The number of trees was evaluated at 50, 100 and 200. Maximum tree depth was evaluated at 10, 20 or unrestricted depth. Square-root feature sampling was used when selecting candidate predictors at individual splits, and bootstrap sampling was enabled.

***Gaussian naïve Bayes (NB).*** Gaussian NB was included as a probabilistic classifier based on the conditional probability of predictor values given class membership. Continuous predictors were modelled using Gaussian class-conditional distributions, with the naïve Bayes assumption that predictors are conditionally independent given class membership. Gaussian NB was implemented using its standard parameter estimation procedure and did not undergo grid-based hyperparameter optimisation.

#### *Training and test procedure*

Predictive performance was evaluated using repeated stratified train-test splitting. At each repetition, 80% of participants were allocated to the training set and the remaining 20% to a held-out test set. Stratification was used to preserve the relative distribution of cognitively impaired and cognitively healthy participants across training and test sets. The complete train-test procedure was repeated 10 times using different random seeds. The initial random state was 42 and was incremented across repetitions, resulting in 10 alternative participant partitions. Critically, the same training and test participant indices were used for the cognitive-only, speech-only and combined predictor sets within each repetition. Consequently, differences in performance between predictor sets within a repetition reflected differences in the information available to the classifier rather than differences in the composition of the test sample. No observations from a held-out test partition were used for model fitting, preprocessing, oversampling or hyperparameter selection.

#### *Missing data and preprocessing*

All preprocessing operations were implemented within the machine-learning pipeline to prevent information leakage from the held-out test data. Missing predictor values were imputed using mean imputation. Importantly, imputation parameters were estimated from the corresponding training data and subsequently applied to the held-out observations rather than being estimated using the complete dataset. Following imputation, predictors were normalised using min-max scaling to transform values to the range 0-1. Scaling parameters were similarly estimated from the training data and subsequently applied to the held-out test data. Normalisation ensured that variables measured on different numerical scales contributed comparably to algorithms sensitive to predictor magnitude. Synthetic Minority Over-sampling Technique (SMOTE) was incorporated into the training pipeline. SMOTE generates synthetic observations within the minority class based on neighbouring observations in feature space. Oversampling was applied only to training data and was not applied to the held-out test set. Because preprocessing and SMOTE were incorporated within the modelling pipeline, these operations were performed separately within the appropriate training folds during cross-validation, preventing synthetic observations or preprocessing information from contaminating validation or test observations. Although the overall analytical sample was approximately balanced between the two outcome classes (58 versus 56), the same preprocessing pipeline was retained consistently across repeated training partitions.

***Hyperparameter optimisation***

Hyperparameter optimisation was conducted using five-fold stratified cross-validation within the training data. The training sample was divided into five folds while preserving class proportions. Models were iteratively trained using four folds and evaluated against the remaining fold, with each fold serving as the validation set once. Area under the receiver operating characteristic curve (AUC) was used as the optimisation criterion. For each train-test repetition, the combination of hyperparameters producing the highest mean cross-validated AUC within the training sample was selected. The resulting optimised model was then evaluated against the held-out test observations. This procedure ensured that model selection and hyperparameter tuning were conducted without reference to the test-set outcomes.

***Model stability and control of overfitting***

Several procedures were implemented to reduce the risk of overfitting. First, all predictor sets were specified according to predefined measurement domains rather than being selected based on test-set performance. Second, all preprocessing, including imputation, scaling and SMOTE, was restricted to the training data and incorporated within the model-fitting pipeline. Third, model hyperparameters were selected using cross-validation within the training sample without access to the held-out test observations. Fourth, performance was assessed on held-out observations and repeated across 10 alternative stratified participant partitions rather than being inferred from training performance or a single test partition. The variability of test-set AUC across repeated partitions was additionally examined as an indicator of model stability. Consistent performance across alternative partitions was considered supportive of model generalisability within the available sample, although the repeated internal validation procedure does not substitute for validation in a fully independent external cohort.

***Evaluation of predictive performance***

Model performance was evaluated exclusively on the held-out test set within each repetition. AUC was designated as the principal measure of discriminatory performance. AUC quantifies the ability of a classifier to discriminate between cognitively impaired and cognitively healthy participants across the full range of possible classification thresholds, with a value of 0.5 representing chance-level discrimination and a value of 1.0 representing perfect discrimination. In addition to AUC, accuracy, precision, recall (sensitivity), specificity and F1 score were calculated. Accuracy represented the overall proportion of correctly classified observations. Precision represented the proportion of participants classified as cognitively impaired who were truly impaired, whereas recall represented the proportion of cognitively impaired participants correctly identified by the classifier. Specificity represented the proportion of cognitively healthy participants correctly classified as healthy. F1 score represented the harmonic mean of precision and recall and was included to provide an overall measure balancing these two aspects of classification performance. Performance metrics were calculated separately for each of the 10 repeated train-test partitions and subsequently summarised using the mean and standard deviation. The repeated-split procedure was used to assess the stability of classification performance across alternative participant partitions and to reduce dependence on a single arbitrary train-test split.

### Supplementary results

The accuracy, sensitivity, specificity and F1 scores for all machine learning models are shown in **Table S1**. Overall, the results indicate that combining cognitive and speech features generally improves classification performance. This pattern was particularly clear for the SVM and RF classifiers, where the combined feature set achieved the highest accuracy, sensitivity, specificity, and F1 scores compared with either feature set alone. Although this pattern was less consistent for NB, where speech features performed better on several metrics, the combined feature set still achieved the highest specificity. Among all models, RF with combined features demonstrated the strongest overall performance, achieving an accuracy of 0.922, sensitivity of 0.908, specificity of 0.936, and F1 score of 0.923.

In secondary analyses comparing the two unimodal feature sets, speech-only models showed higher AUCs than cognitive-only models across all three classifiers. The difference was statistically significant for NB ($\Delta AUC = .069$, $p = .036$) and RF ($\Delta AUC = .073$, $p = .015$), but not for SVM ($\Delta AUC = .059$, $p = .069$). These findings indicate that speech-derived features generally provided greater discriminatory performance than cognitive features alone, although statistical evidence for this difference was not observed across all classification models.

### Supplementary table

| Classifier | Feature set | Accuracy | Sensitivity | Specificity | F1 |
|---|---|---|---|---|---|
| NB | Cognitive | .817 | .658 | .991 | .783 |
| NB | Speech | .878 | .817 | .945 | .872 |
| NB | Combined | .865 | .750 | .991 | .846 |
| SVM | Cognitive | .839 | .742 | .945 | .819 |
| SVM | Speech | .852 | .817 | .891 | .850 |
| SVM | Combined | .900 | .833 | .973 | .891 |
| RF | Cognitive | .804 | .708 | .909 | .789 |
| RF | Speech | .870 | .842 | .900 | .869 |
| RF | Combined | .922 | .908 | .936 | .923 |

**Table S1: A table showing the accuracy, sensitivity, specificity and F1 scores for all machine learning models.** *Abbreviations:* NB=naïve bayes; SVM=support vector machine; RF=random forest
